# Transmission Waiting Time: A Unifying Metric for Outbreak Controllability

**DOI:** 10.1101/2025.10.18.25338292

**Authors:** Christopher Y. Shin, Sang Woo Park, Cécile Viboud, Pardis C. Sabeti, Christophe Fraser, Kaiyuan Sun

## Abstract

The feasibility of containing infectious disease outbreaks depends on whether interventions can act faster than pathogen transmission. Classical controllability theory links outbreak control to the timing of symptom onset relative to infectiousness, reflecting an era in which isolation and contact tracing were necessarily triggered by clinical symptoms. This framework therefore implies intrinsic limits for pathogens with substantial pre-symptomatic or asymptomatic transmission. However, the widespread availability of molecular diagnostics now enables infection detection and isolation independent of symptoms, fundamentally altering the operational basis of outbreak control. Here we introduce the transmission waiting time—the interval between infection and the first onward transmission event—as a symptom-agnostic timescale that defines the intrinsic speed limit for effective intervention. We derive this quantity analytically from two fundamental epidemiological parameters, the basic reproduction number and the generation-interval distribution, yielding closed-form benchmarks for the minimum isolation speed and coverage required for control. Applying this framework across diverse pathogens reveals substantial differences in intrinsic controllability driven by variation in both overall transmissibility and the timing of infectiousness. We show that transmission heterogeneity reshapes controllability primarily through changes in the temporal distribution of infectiousness, and that homogeneous predictions often provide conservative bounds. By mapping complex test–trace–isolate operations onto a low-dimensional delay–coverage intervention landscape, this framework clarifies the conditions under which such containment efforts are feasible.

## Main

The controllability of infectious disease outbreaks is determined by a race between pathogen transmission and the speed of public health intervention. ^1,2^ For decades, this race was governed by a symptom-based paradigm, in which isolation and contact tracing were initiated only after the onset of clinical signs. Formalized following the 2003 SARS-CoV-1 epidemic, classical theory framed outbreak controllability in terms of a pathogen’s basic reproduction number, *R*_0_, and the fraction of transmission occurring before symptom onset. ^1^ This framework explains the successful containment of pathogens such as SARS-CoV-1 and smallpox—where symptoms reliably precede infectiousness—and the intrinsic difficulty of controlling pathogens such as influenza, where substantial transmission occurs before symptoms. ^3–5^ Reliance on symptom-triggered response imposes a structural delay on the order of the incubation period, rendering containment infeasible when transmission is largely pre-symptomatic or when infections are frequently asymptomatic.

SARS-CoV-2 emerged in a fundamentally different intervention landscape: its extensive presymptomatic transmission rendered symptom-based control too slow, while the widespread availability of molecular diagnostics enabled infection detection independent of symptom presentation. ^6–11^ This shift moved the bottleneck for outbreak control from clinical recognition to operational constraints—such as testing frequency, diagnostic turnaround time, notification delay, and the timeliness of isolation—that jointly determine intervention coverage and delay. Concurrent technological advances, including smartphone-based digital contact tracing and rapid, high-sensitivity point-of-care diagnostics, further demonstrated the potential to shorten detection-to-notification intervals and accelerate response. ^12–19^ Yet, despite these advances, a unifying theory of outbreak controllability that links operational intervention delays to the intrinsic temporal constraints imposed by pathogen biology remains lacking.

### Transmission waiting time: the critical timescale for outbreak control

To address this gap, we introduce a theoretical framework centered on the transmission waiting time, *w*(*τ* ): the interval between infection and the first onward transmission event. In contrast to the classical concept of generation interval distribution *g*(*τ* )—which describes the timing of all secondary transmissions—*w*(*τ* ) isolates the earliest transmission event, the moment an intervention must pre-empt to block onward spread.

Failure to pre-empt this earliest transmission event leaves a substantial opportunity for generating secondary infections and sustaining transmission chains, rendering outbreak control infeasible. The transmission waiting time therefore defines an intrinsic speed limit for epidemic containment. This intuition constitutes the central principle of the manuscript and motivates the development of a rigorous theoretical framework for pathogen controllability in the sections that follow.

Starting from a homogeneous population, we derive *w*(*τ* ) analytically from two fundamental epidemiological quantities of a pathogen: the basic reproduction number *R*_0_, which encodes a pathogen’s overall transmissibility, and the generation interval distribution *g*(*τ* ), which characterizes the average temporal profile of infectiousness over the course of an infection episode. Under this framework, secondary transmissions arise according to a non-stationary Poisson process with transmission hazard *h*(*τ* ) = *R*_0_ *g*(*τ* ). Using standard survival-analysis techniques (Methods), the transmission waiting time distribution, conditional on at least one secondary transmission occur-ring, is given by

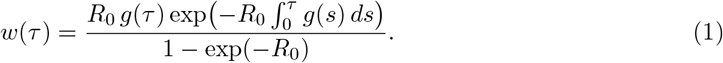

This expression shows that the timing of the first transmission event is fully determined by *R*_0_ and *g*(*τ* ).

We next examine how the transmission waiting time constrains isolation-based interventions. Although such interventions vary widely in implementation—from symptom-driven isolation and manual contact tracing to routine rapid-test screening and digital exposure notification^20–22^—their epidemiological effects can be summarized by two operational quantities: the isolation coverage, *P*_iso_, defined as the fraction of infections that are ultimately identified and isolated, and the isolation-delay distribution, *δ*_iso_(*τ* ), which characterizes the timing of isolation among those individuals. The probability that an infected individual remains unisolated at time *τ* is therefore

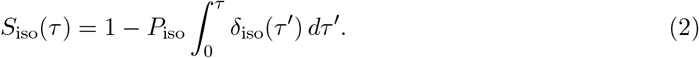

Subsequently, the corresponding effective reproduction number under intervention is given by

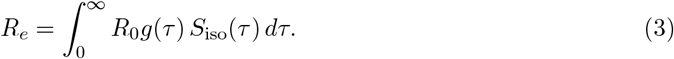

Under perfect coverage (*P*_iso_ = 1), if isolation delays exactly match the transmission waiting time 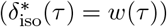, the effective reproduction number reduces to

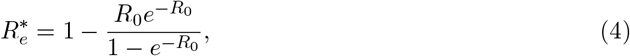

which is strictly less than one for all *R*_0_ and approaches unity only asymptotically as *R*_0_ becomes large (Methods). Thus, any intervention that acts at least as rapidly as the intrinsic transmission waiting time is, in principle, sufficient to guarantee outbreak control.

Because *w*(*τ* ) is defined conditional on at least one secondary transmission, it integrates to unity even when the probability that an infection generates no onward transmission—given by 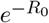 —is substantial for relatively small values of *R*_0_. In this low *R*_0_ regime, requiring isolation delays to exactly match *w*(*τ* ) under perfect isolation coverage is therefore unnecessarily stringent. Relaxing this constraint, we instead impose *R*_*e*_ = 1 while retaining 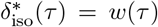, thereby yielding the critical isolation coverage (Methods).

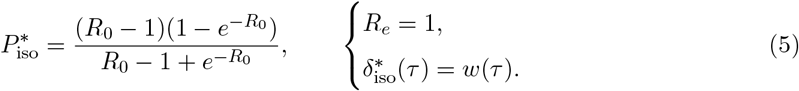

This quantity depends solely on *R*_0_ and increases monotonically toward unity as transmissibility rises, indicating that pathogens with large *R*_0_ require isolation of nearly all infectious individuals—even when isolation operates at the speed defined by the transmission waiting time. Together, *w*(*τ* ) and 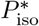 delineate the minimal speed and reach of isolation required for containment, directly linking the feasibility of outbreak control to a pathogen’s intrinsic transmissibility and temporal infectiousness profile.

### Epidemiological determinants of pathogen controllability

To assess the practical utility of the transmission waiting time framework, we first examined how waiting times depend on fundamental epidemiological parameters. Stochastic ordering analysis (Methods) shows that transmission waiting times become systematically shorter when transmission is more intense (higher *R*_0_; Fig. 1A) and when infectiousness is concentrated earlier in infection (shorter generation intervals; Fig. 1B). Together, these results formalize the intuitive notion that both the overall magnitude and the temporal placement of infectiousness determine the intrinsic speed of transmission.

**Figure 1.**
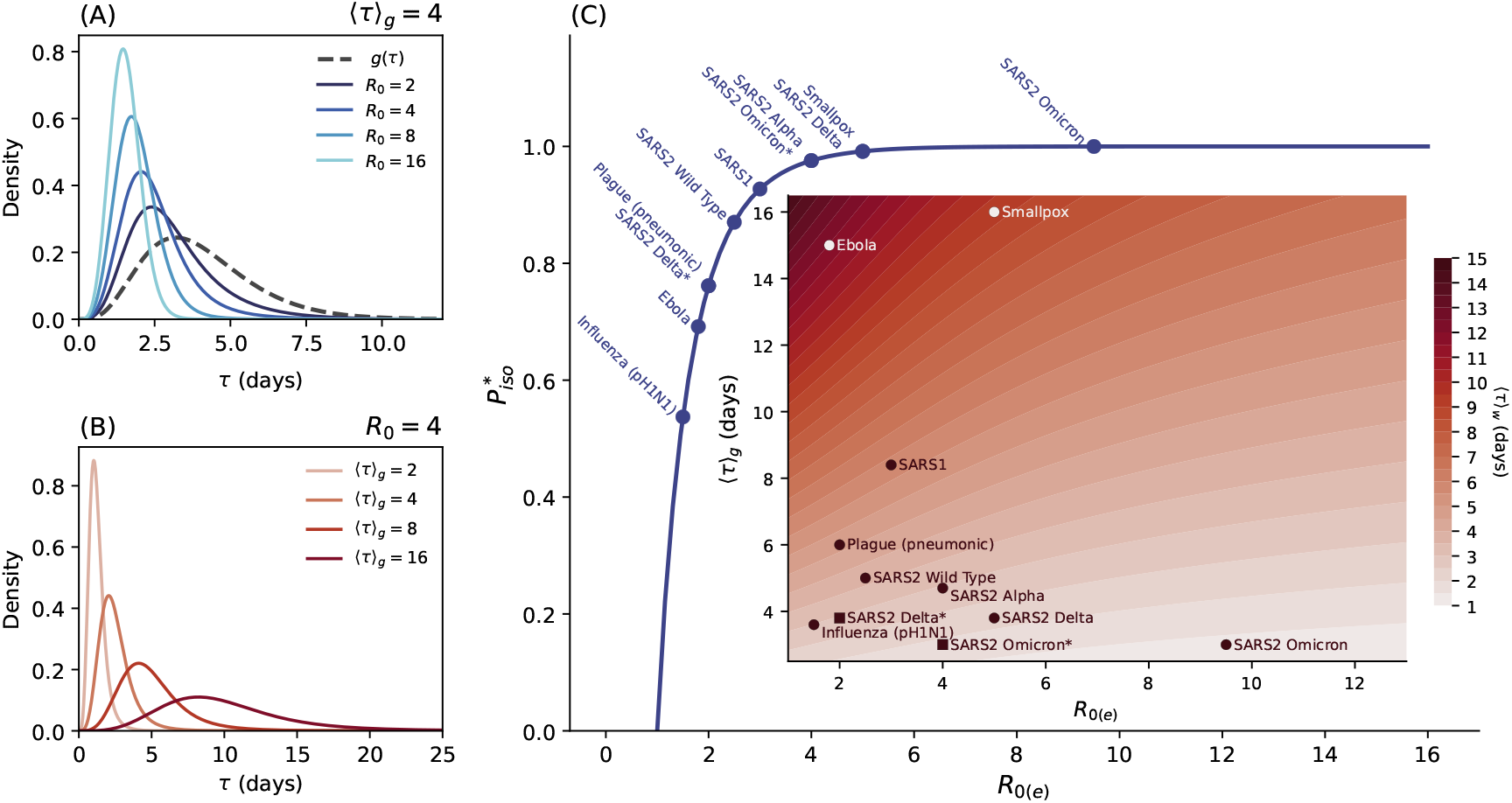
Epidemiological determinants of transmission waiting time and isolation requirements. (A) Effect of increasing the basic reproduction number *R*_0_ on the transmission waiting time distribution *w*(*τ* ), holding the generation-interval distribution *g*(*τ* ) fixed (gamma-distributed with mean 4 days and shape parameter *β* = 5). The corresponding generation-interval distribution *g*(*τ* ) is shown as a dashed curve. Larger *R*_0_ shifts *w*(*τ* ) toward earlier times, shortening the effective window for intervention. (B) Effect of decreasing the mean generation interval ⟨*τ*⟩_*g*_ on *w*(*τ* ), holding *R*_0_ = 4 and *β* = 5 fixed. Shorter generation intervals concentrate transmission earlier during infection, leading to similarly reduced waiting times. (C) Main panel: Relationship between the basic reproduction number *R*_0_ and the minimum isolation coverage 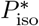 required for containment (*R*_*e*_ = 1). Points denote empirical estimates for selected pathogens, while the solid curve shows the theoretical prediction. Circle markers indicate basic reproduction numbers, and square markers indicate effective reproduction numbers. Inset: Heatmap showing how the mean transmission waiting time ⟨*τ*⟩_*w*_ varies jointly with *R*_0_ and the mean generation interval, highlighting multiple epidemiological pathways to rapid transmission. SARS1: SARS-CoV-1; SARS2: SARS-CoV-2; Delta*: effective reproduction number of the SARS-CoV-2 Delta variant reduced to 2 under non-pharmaceutical interventions; Omicron*: effective reproduction number of the SARS-CoV-2 Omicron variant reduced to 4 due to prior immunity.

We next applied the transmission waiting time framework to a representative set of pathogens that spread predominantly through direct person-to-person contact. For each pathogen, we compiled published estimates of the basic reproduction number *R*_0_ and the generation-interval distribution *g*(*τ* ). In addition, for selected SARS-CoV-2 variants, we also considered scenarios in which non-pharmaceutical interventions or prior immunity reduce transmission, and therefore report the effective reproduction number *R*_*e*_ instead of *R*_0_. These inputs were used to derive the corresponding mean transmission waiting time, ⟨*τ*⟩_*w*_, together with the minimum isolation coverage required for containment, 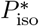 . Estimates of *R*_0_ (or *R*_*e*_, where applicable), the mean generation interval ⟨*τ*⟩_*g*_, the mean transmission waiting time ⟨*τ*⟩_*w*_, and the critical isolation threshold 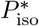 for all pathogens considered are summarized in Table 1.

**Table 1:** Epidemiological parameters and critical isolation requirements for selected pathogens. *indicates effective reproduction number *R*_*e*_ instead of basic reproduction number *R*_0_ is reported. NPI: Non-pharmaceutical intervention.

| Pathogen | $R_{0(e)}$ | $\langle\tau\rangle_g$ | $\langle\tau\rangle_w$ | $P_{\text{iso}}$ | Reference(s) |
| --- | --- | --- | --- | --- | --- |
| Ebola | 1.8 | 15.0 | 11.96 | 0.69 | 23,24 |
| Smallpox | 5.0 | 16.0 | 9.24 | 0.99 | 3,25,26 |
| SARS-CoV-1 | 3.0 | 8.4 | 5.83 | 0.93 | 1,27 |
| Plague (pneumonic) | 2.0 | 6.0 | 4.61 | 0.76 | 28,29 |
| SARS-CoV-2 (Wild Type) | 2.5 | 5.0 | 3.67 | 0.87 | 30,31 |
| 2009 H1N1 pandemic | 1.5 | 3.6 | 2.98 | 0.54 | 32,33 |
| SARS-CoV-2 (Delta*) | 2.0 | 3.8 | 2.96 | 0.76 | *$R_e = 2.0$ (under NPI) |
| SARS-CoV-2 (Alpha) | 4.0 | 4.7 | 2.95 | 0.98 | 34 |
| SARS-CoV-2 (Delta) | 5.0 | 3.8 | 2.19 | 0.99 | 35,36 |
| SARS-CoV-2 (Omicron*) | 4.0 | 3.0 | 1.88 | 0.98 | *$R_e = 4.0$ (prior immunity) |
| SARS-CoV-2 (Omicron) | 9.5 | 3.0 | 1.37 | 1.00 | 35,37–39 |
| Unit | – | days | days | – | – |

Figure 1C visualizes these dependencies across the joint parameter space of *R*_0_ and the mean generation interval, highlighting that rapid transmission can arise through multiple, non-mutually exclusive mechanisms: high transmissibility, rapid inter-generational turnover, or both. This comparative analysis reveals substantial variation in intrinsic controllability across pathogens.

Pathogens such as Ebola virus, which combine long generation intervals with moderate transmissibility, exhibit long transmission waiting times (approximately 12 days), providing relatively wide temporal windows for effective intervention. ^23,24^ Smallpox, despite having a generation interval of comparable length, is characterized by substantially higher transmissibility and therefore shorter transmission waiting times, requiring near-perfect isolation coverage and rendering containment operationally more challenging. ^3,25,26^ In contrast, the 2009 pandemic H1N1 influenza strain has a very short generation interval and consequently exhibits transmission waiting times of only 2–3 days even at modest values of *R*_0_, illustrating that rapid inter-generational turnover alone can significantly constrain the feasibility of outbreak control. ^32,33^

The evolutionary trajectory of SARS-CoV-2 further underscores the central role of the transmission waiting time in shaping pathogen controllability. The ancestral strain (wild type) exhibited intermediate waiting times (approximately 3–4 days), consistent with the transient containment achieved in several regions through early test–trace–isolate strategies.^30,31^ However, successive variants of concern evolved toward both higher transmissibility and shorter generation intervals, resulting in progressively shorter transmission waiting times ⟨*τ*⟩_*w*_—a trend most clearly illustrated by the emergence of the Omicron variant.^34–39^ With a transmission waiting time of roughly 1.37 days and a critical isolation coverage approaching unity, Omicron effectively placed isolation-based containment beyond operational reach in most settings. ^35,37–39^

Importantly, both population-level non-pharmaceutical interventions (e.g., mobility restrictions) and accumulated population immunity can reduce the effective reproduction number and extend transmission waiting time. As shown in Table 1 and Fig. 1C, scenarios corresponding to the Delta variant under intervention and the Omicron variant in populations with substantial prior immunity exhibit longer transmission waiting times compared with their uninhibited spread in immunologically naïve populations. These comparisons highlight how reductions in effective transmissibility can meaningfully shift the transmission timescale, even when intrinsic viral properties favor rapid spread.

### Controllability in the presence of transmission heterogeneity

The preceding analysis established outbreak controllability under the homogeneous assumption, in which all infected individuals share a common reproduction number *R*_0_ and generation-interval distribution *g*(*τ* ). Real-world populations, however, exhibit substantial heterogeneity in pathogen transmission. Infected individuals vary both in the number of secondary infections they generate and in the timing of infectiousness, and these two dimensions of heterogeneity are likely to be correlated. ^40–42^ A central question, therefore, is whether such heterogeneity qualitatively alters the conclusions derived under homogeneous assumptions, or whether it primarily manifests as a structured perturbation around that benchmark.

To address this question, we consider a population in which each infected individual is characterized by two latent attributes: an individual transmissibility parameter *R*, defined as the individual reproduction number, and an individual timing of infectiousness parameter *µ*_*g*_, defined as the mean of the individual generation-interval distribution *g*(*τ* | *µ*_*g*_). Under this formulation, the individual-level transmission hazard is given by

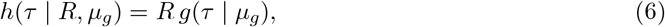

where 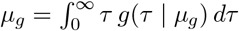.

For a given pathogen, it is biologically plausible that *R* increases with *µ*_*g*_, such that individuals who has longer mean generation interval tend, on average, to generate more secondary infections. To capture this dependence, we model the individual reproduction number as

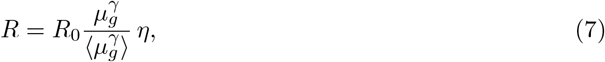

where *µ*_*g*_ follows a population-level distribution *f*_*g*_(*µ*_*g*_), reflecting heterogeneity in the timing of infectiousness. The parameter *γ ∈* [0, 2] governs the strength and form of the coupling between *R* and *µ*_*g*_: *γ* = 0 corresponds to no dependence; *γ* = 1 implies linear scaling of *R* with *µ*_*g*_; and *γ* = 2 represents linear scaling with total infectiousness over time, corresponding to the area under an individual infectiousness profile when both shedding intensity and duration increase with *µ*_*g*_. The random variable *η* captures residual, *µ*_*g*_-independent heterogeneity in transmissibility and its distribution *f*_*η*_(*η*) is assumed to be non-negative with unit mean 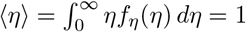 . Together with the normalization factor 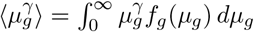 this formulation ensures that the population average of individual reproduction number ⟨*R*⟩ = *R*_0_ for all choices of *{f*_*g*_, *γ, f*_*η*_*}*.

Because individual-level transmission attributes are generally unobservable in real time, interventions targeted to individual risk are impractical in most settings. We therefore consider uniform isolation strategies, characterized by a single isolation coverage *P*_iso_ and a shared isolation-delay distribution *δ*_iso_(*τ* ), applied identically across the population. Under this assumption, the probability that an infected individual remains unisolated at infection age *τ* is the same as in the homogeneous-mixing case: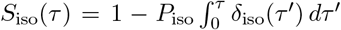 . The population-average effective reproduction number is then given by the expected number of secondary infections generated before isolation, averaged over the joint distribution of individual attributes *µ*_*g*_ and *η*:

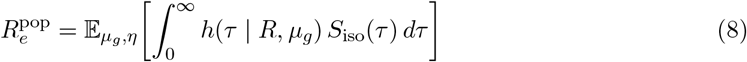

Notably, the residual heterogeneity term *η*, which captures variation in transmissibility independent of infectiousness timing, integrates out and does not affect the population-level effective reproduction number:

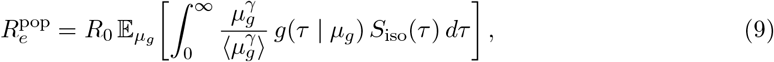

which implies 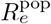 depends only on heterogeneity in infectiousness timing and its correlation with transmissibility. Consequently, heterogeneity in transmissibility alone does not alter outbreak controllability at the population level; only heterogeneity that redistributes infectiousness in time—and its coupling to individual transmissibility *R*—can shift the isolation requirements relative to those predicted under homogeneous mixing.

### The impact of heterogeneity in *R* on controllability under homogeneous *µ*_*g*_

To elaborate on the point above, we first consider a limiting case in which transmission heterogeneity arises solely from variation in individual transmissibility *R*, while infectiousness timing is homogeneous across individuals. Specifically, the residual heterogeneity term *η* follows a gamma distribution with shape parameter *k*, and all individuals share an identical generation-interval distribution *g*(*τ* ), implying no variation in *µ*_*g*_.

Under these assumptions and in the absence of intervention, the offspring distribution is negative binomial with mean *R*_0_ and dispersion parameter *k*, producing substantial variation in individual secondary infection counts when *k* is small. Nevertheless, because the population-level effective reproduction number *R*_*e*_ does not depend on *η* (Eq. (9)), and infectiousness timing is identical across individuals, the expression for *R*_*e*_ under intervention reduces exactly to the homogeneous-mixing result (Eq. (3)). Consequently, under the assumption of a homogeneous generation-interval distribution, variation in individual reproduction numbers—although it induces pronounced overdispersion in the distribution of secondary infections—does not modify the outbreak controllability when compared with the corresponding homogeneous reference scenario.

### The impact of heterogeneity in infectiousness timing on controllability

To quantify how heterogeneity in infectiousness timing reshapes outbreak controllability, we consider a stylized pathogen with basic reproduction number *R*_0_ = 3. Individual infectiousness timing parameter *µ*_*g*_ is drawn from a gamma distribution with mean ⟨*µ*_*g*_⟩ = 6 days and shape parameter *α*, such that smaller *α* corresponds to greater heterogeneity, while *α →* ∞ recovers the homogeneous-mixing limit. Conditional on *µ*_*g*_, the individual generation interval distribution *g*(*τ* | *µ*_*g*_) is gamma-distributed with fixed shape parameter and individual-specific mean (*µ*_*g*_).

We allow individual transmissibility to scale with infectiousness timing according to 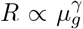, where *γ* controls the strength of coupling. All results assume uniform isolation with perfect coverage (*P*_iso_ = 100%) and vary only the mean isolation delay ⟨*τ*⟩_iso_ (detailed in Methods).

Figure 2 illustrates how heterogeneity in *µ*_*g*_, and its correlation with transmissibility, alters the isolation speed required for control. The inset panels show the population-average effective reproduction number 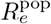 as a function of ⟨*τ*⟩_iso_ for two limiting cases. When transmissibility is independent of infectiousness timing (*γ* = 0), increasing heterogeneity raises 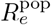 for a given isolation delay, requiring faster isolation than predicted under homogeneous mixing. This tightening reflects the increased contribution of early transmission from individuals with short generation intervals, which are difficult to pre-empt using a uniform intervention.

**Figure 2.**
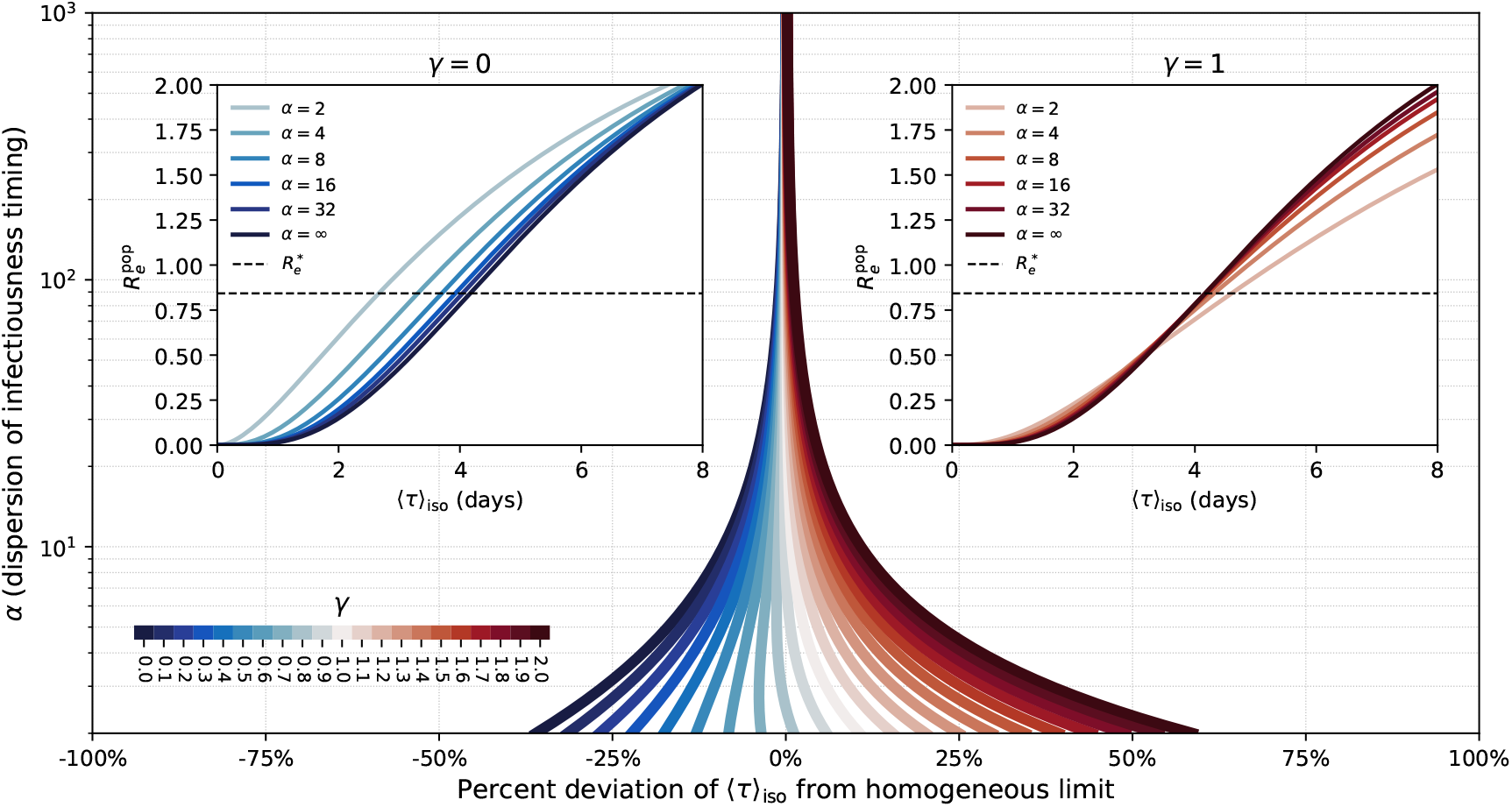
Heterogeneity in infectiousness timing modifies outbreak controllability. Inset panels show the population-average effective reproduction number 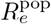 as a function of the mean isolation delay ⟨*τ*⟩_iso_ for varying heterogeneity in infectiousness timing (*α*) under two correlation strengths between transmissibility and duration (*γ* = 0 and *γ* = 1). The horizontal dashed line denotes the critical threshold 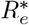, defined by the homogeneous transmission waiting time prediction under perfect isolation coverage (Eq. (4)). The main panel shows the percentage deviation in the isolation delay required to achieve 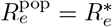 relative to the homogeneous-mixing benchmark.

By contrast, when transmissibility increases with mean generation interval (*γ* = 1), these effects are strongly attenuated. Individuals with shorter *µ*_*g*_ also generate fewer secondary infections, partially offsetting their earlier transmission. As a result, isolation requirements remain close to the homogeneous-mixing prediction across a wide range of heterogeneity.

The main panel summarizes these effects by plotting, across values of *α* and *γ*, the percentage deviation in the mean isolation delay required to achieve 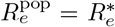 relative to the homogeneous benchmark. Deviations vanish as *α →* ∞, but become increasingly pronounced at low *α*. Increasing correlation strength progressively shifts these deviations toward zero and, under strong coupling, can even reverse their sign, indicating that heterogeneity may relax isolation-speed requirements when transmission is concentrated among individuals with longer, more interceptable infectiousness profiles.

Taken together, these results show that introducing heterogeneity does not overturn the qualitative conclusions of the transmission waiting time framework; instead, it acts as a structured perturbation around the homogeneous limit. Given empirical evidence that infectiousness timing and overall shedding intensity are often positively correlated, the homogeneous theory therefore provides a conservative bound on outbreak controllability. Whether heterogeneity ultimately tightens or relaxes the speed requirements for containment is governed by how transmission potential is redistributed in time relative to intervention, rather than by variation in the individual reproduction number alone.

### Operational determinants of pathogen controllability

The transmission waiting time framework links outbreak controllability to two operational quantities: the fraction of infections that are successfully isolated (isolation coverage) and the timing of isolation relative to infectiousness profile (isolation delay). A broad range of implementation-specific factors—including symptom recognition, testing frequency, diagnostic sensitivity, turnaround time, notification speed, contact tracing efficiency, and adherence—primarily influence pathogen controllability through their effects on these two quantities. We therefore evaluate intervention performance through the joint lens of isolation coverage and isolation delay, first using a stylized comparison between symptom-based and testing-based control to illustrate how advances in diagnostic technologies reshape intervention performance, and then embedding these insights within a more realistic intervention landscape using agent-based simulations.

Under the symptom-based intervention paradigm, isolation is initiated upon the onset of clinical symptoms. Consequently, the delay between infection and intervention is determined by the distribution of the incubation period—the time interval between infection and symptom onset—among individuals who develop symptoms. For pathogens with substantial pre-symptomatic transmission, this delay is structurally constrained to occur after a large fraction of transmission has already taken place. ^7^ Using SARS-CoV-2 as a case study, Fig. 3(A) compares incubation-period distributions with variant-specific transmission waiting times. From the ancestral lineage through Omicron, the mean incubation period consistently exceeds the corresponding transmission waiting time, indicating that symptom-triggered isolation systematically lags behind the earliest transmission events. The presence of a substantial fraction of asymptomatic infections further erodes effectiveness, rendering symptom-based containment infeasible for SARS-CoV-2. This limitation contrasts sharply with SARS-CoV-1. For SARS-CoV-1, the incubation period is shorter than the transmission waiting time (Fig. 3(A), inset), reflecting limited pre-symptomatic transmission. Combined with its high symptomatic rate, this temporal ordering—where symptom onset reliably precedes significant onward transmission—enabled symptom-based isolation and contact tracing to effectively interrupt transmission chains and achieve outbreak control, in alignment with the classical controllability theory established after the 2003 SARS outbreak. ^1^

**Figure 3.**
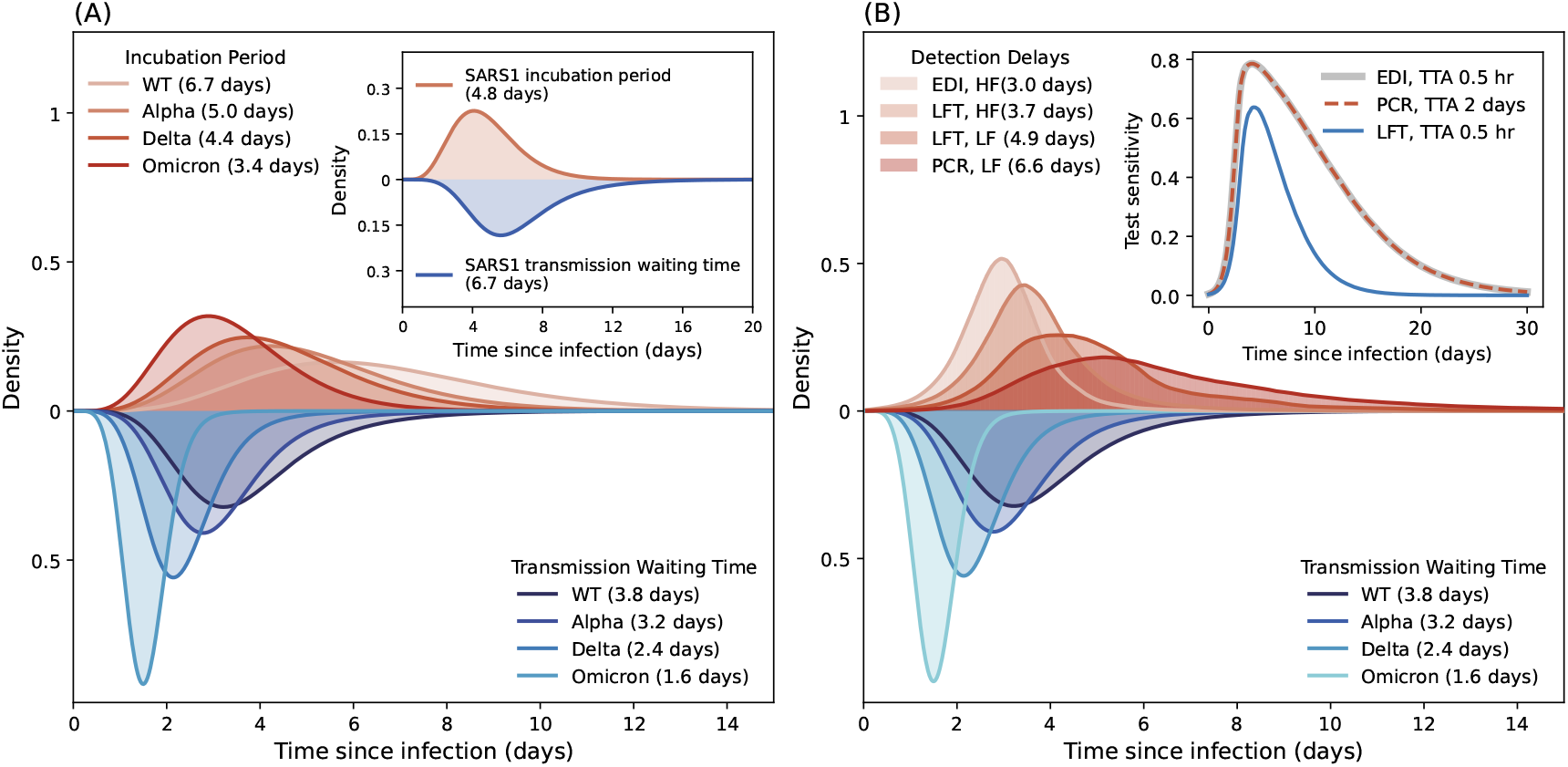
Pathogen controllability through symptom-driven versus testing-driven isolation-based interventions. **(A)** Distributions of incubation periods (symptom-based isolation delays) for SARS-CoV-2 variants (wild type (WT), Alpha, Delta, Omicron) compared with their corresponding transmission waiting times (means indicated in parentheses). Inset panel: distribution of incubation period and transmission waiting time for SARS-CoV-1 (SARS1, means indicated in parentheses). **(B)** Distributions of infection detection delays under population-wide testing strategies, compared with variant-specific transmission waiting times (means indicated in parentheses). We consider three diagnostic modalities: (1) PCR targeting viral RNA; (2) LFT targeting viral antigen; and (3) EDI that combine PCR-like sensitivity with LFT-like point-of-care speed. Test sensitivity as a function of time since infection as will as the testing turnaround (TTA) are reported in the inset panel. the Detection delays are shown for PCR and LFT under low-frequency (LF) testing, and for LFT and EDI under high-frequency (HF) testing.

Testing-based interventions overcome the structural delay imposed by symptom onset through decoupling case detection from clinical presentation, enabling identification of infections regardless of symptom status. To illustrate this, we first consider a stylized test-and-isolate intervention: individuals are tested at regular intervals regardless of symptoms and immediately isolate upon receiving a positive result. The resulting intervention delay is governed by three components: the earliest time at which infection becomes detectable, determined by the assay’s limit of detection relative to viral kinetics; the testing frequency; and the diagnostic turnaround time from sample collection to outcome notification. ^10^ Conventional diagnostic technologies involve a trade-off between sensitivity and speed: reverse transcription–polymerase chain reaction assays (RT–PCR, referred to hereafter as PCR) can detect low viral loads early in infection but often entail significant delays due to centralized laboratory processing, whereas rapid antigen lateral flow test (LFT) pro-vide near-immediate, point-of-care results yet typically identify infections only at later stages and higher viral loads, owing to their lower analytical sensitivity. Emerging molecular diagnostic innovation (EDI)—including CRISPR-based assays, isothermal amplification platforms, and portable microfluidic PCR systems—aim to combine high sensitivity with rapid turnaround. ^43–49^

Figure 3(B) compares the distributions of isolation delays under population-wide testing strategies against variant-specific transmission waiting times. We assume a testing turnaround time of 2 days for PCR and 0.5 hours for both LFT and EDI. Two testing frequency scenarios are considered: a high-frequency (HF) scenario, in which the entire population is tested daily, and a low-frequency (LF) scenario, with population tested every three days. Test sensitivity as a function of time since infection is drawn from published literature for both PCR and LFT; for EDI, sensitivity is assumed to match that of PCR.^50^

Under the LF scenario, mean isolation delays exceed the transmission waiting time of the ancestral SARS-CoV-2 lineage (mean = 3.8 days)—6.6 days for PCR and 4.9 days for LFTs. Despite its higher sensitivity, PCR incurs longer delays due to its extended turnaround time (2 days vs. 0.5 hours). In contrast, the HF scenario—especially when combined with rapid-turnaround assays such as LFT and EDI—substantially compresses isolation delays to levels comparable to or shorter than the transmission waiting times of early SARS-CoV-2 variants. High-frequency LFT testing reduces the mean detection delay to 3.7 days, while EDI further shorten it to approximately 3.0 days—sufficient to control early variants like Alpha without additional contact tracing or population level travel restrictions. Across all strategies evaluated, fewer than 10% of infections were missed, reflecting high intervention coverage due to effective infection ascertainment.

Population-wide access to routine molecular testing can shift the distribution of isolation delays toward—and in some cases ahead of—the intrinsic transmission timescale that defines pathogen controllability within our framework. In practice, however, outbreak control rarely relies on testing alone. Instead, control is typically achieved through layered intervention systems that combine symptom-based surveillance, population-level mobility restrictions, routine diagnostic screening, and contact tracing, each operating under logistical constraints that introduce delays and limit coverage. As a result, real-world intervention systems are substantially more complex than the idealized scenarios considered above. Their effectiveness is constrained by operational limitations, such as finite testing capacity, diagnostic turnaround delays, incomplete contact tracing, and imperfect adherence. These factors jointly shape the performance of non-pharmaceutical interventions (NPIs), particularly test–trace–isolate (TTI) strategies.^20–22^ To bridge the gap between the theoretical controllability thresholds defined by the transmission waiting time framework and realistic outbreak-control implementation, we therefore conducted a simulation-based intervention analysis using a branching process model.

Specifically, we simulated a hypothetical respiratory pathogen with a basic reproduction number *R*_0_ = 6 and a mean generation interval of 6 days. We assumed the presence of background population-level interventions—such as travel restrictions—that reduced the effective reproduction number to 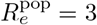 prior to the implementation of TTI interventions. Against this population-wide intervention backdrop, we evaluated six TTI scenarios, each representing incremental improvements in symptom-based isolation compliance, testing frequency and sensitivity, diagnostic turnaround time, and contact-tracing delay (Fig. 4, inset table). To connect these detailed operational components to our theoretical framework, we summarize intervention performance using two aggregate quantities: the mean isolation delay, ⟨*τ*⟩_iso_, which captures the timeliness of isolation relative to infection, and the isolation coverage, *P*_iso_, which represents the fraction of infections successfully isolated. Collectively, these scenarios span a realistic range of operational performance observed in practice. Full model details are provided in the Methods.

**Figure 4.**
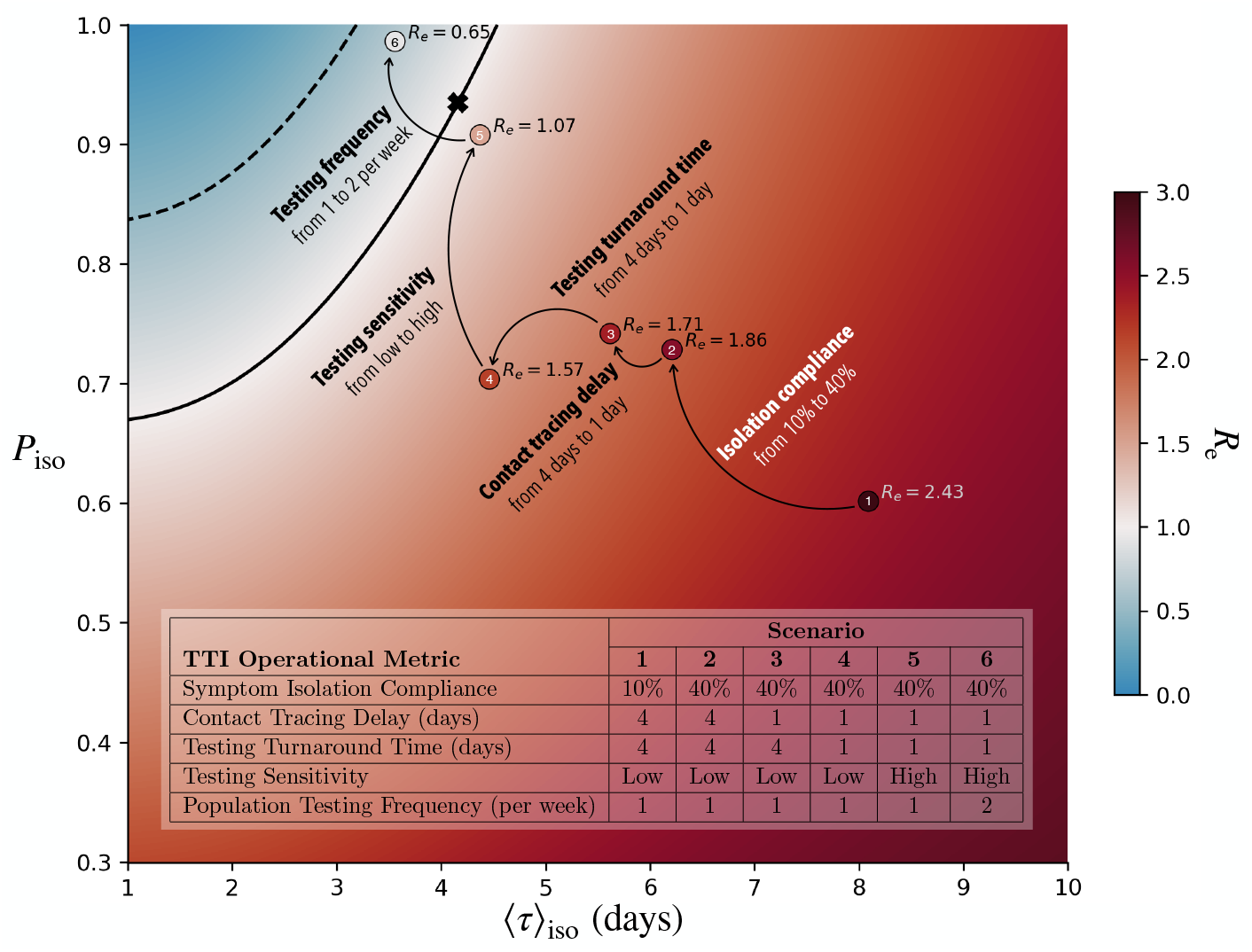
Intervention landscape linking isolation timeliness and coverage to outbreak controllability under test–trace–isolate (TTI) strategies. The heatmap shows the effective reproduction number *R*_*e*_ as a function of the mean isolation delay ⟨*τ*⟩_iso_ and isolation coverage *P*_iso_, assuming moderate background population-level mitigation. Contour lines indicate the theoretical containment boundary (*R*_*e*_ = 1) in the absence of both population-level and case-based interventions (*R*_0_ = 6, dashed line) and under population-level control alone ( 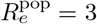, solid line). Conditional on background population-level measures, colored markers denote simulated TTI scenarios with incremental improvements in operational performance, while the black cross marks the theoretical control condition at which the intervention timescale coincides with the transmission waiting time, ⟨*τ*⟩_iso_ = ⟨*τ*⟩_*w*_. **Inset table:** summary of the six incremental TTI scenarios.

Figure 4 situates these scenarios within an intervention landscape defined by isolation delay and isolation coverage. The heatmap shows the effective reproduction number *R*_*e*_ as a function of ( ⟨*τ*⟩_iso_, *P*_iso_). Contour lines denote the theoretical containment boundary (*R*_*e*_ = 1) under moderate population-level mitigation (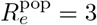, solid line) and in the absence of population-wide mitigation (*R*_0_ = 6, dashed line). Conditional on the assumed background population-level control, colored markers indicate the simulated test–trace–isolate (TTI) scenarios, while the black cross marks the theoretical control condition at which the mean isolation delay coincides with the transmission waiting time, ⟨*τ*⟩_iso_ = ⟨*τ*⟩_*w*_.

As operational performance improves—through reductions in isolation delay and/or increases in isolation coverage—scenarios move systematically across the landscape toward the containment boundary, accompanied by progressively lower effective reproduction numbers. The most effective configuration (Scenario 6), which combines high testing frequency, rapid diagnostic turnaround, efficient contact tracing, and background population-level mitigation, crosses this boundary and achieves *R*_*e*_ *<* 1.

Together, these results underscore the essential and complementary roles of intervention speed and coverage in outbreak control. By projecting complex, multi-component intervention systems onto a low-dimensional delay–coverage plane, the transmission waiting time framework provides a quantitative bridge between operational constraints and intrinsic controllability timescales. This representation offers a principled basis for comparing, evaluating, and optimizing TTI strategies under realistic implementation conditions.

## Discussion

Over the past two decades, the operational determinants of outbreak control have shifted fundamentally. During the 2003 SARS-CoV-1 epidemic, containment relied primarily on symptom-based case detection and contact tracing. In contrast, the extensive pre-symptomatic and asymptomatic transmission of SARS-CoV-2 rendered symptom-triggered interventions structurally too slow, even when implemented at scale. ^1,6–11^ At the same time, widespread molecular diagnostics, routine screening, and digital exposure notification enabled symptom-independent identification and isolation, shifting the effective bottleneck from clinical recognition to operational performance—including testing frequency, turnaround time, notification delays, tracing efficiency, and adherence. ^12–19,51^ Motivated by this transition, we developed a symptom-independent theory of outbreak controllability that compares the timing of interventions directly against the earliest opportunities for transmission.

Our framework generalizes the influential paradigm introduced by Fraser et al., which linked controllability to the basic reproduction number *R*_0_ and the timing of symptom onset relative to infectiousness. ^1^ Whereas classical theory emphasizes the structural delay imposed by symptom-based surveillance, we anchor containment to a more fundamental temporal constraint: the first onward transmission event. We formalize this constraint through the transmission waiting time, *w*(*τ* ), defined as the interval between infection and the first secondary transmission, conditional on at least one transmission occurring. Because failure to pre-empt this first event leaves substantial remaining opportunity for onward spread, *w*(*τ* ) defines an intrinsic speed limit for isolation-based containment. Under homogeneous transmission, we show that *w*(*τ* ) is fully determined by *R*_0_ and the generation-interval distribution *g*(*τ* ), yielding closed-form benchmarks for both the critical isolation-delay distribution and the minimum isolation coverage required to achieve *R*_*e*_ *≤* 1. Because *R*_0_ and *g*(*τ* ) can often be estimated at the early phase of an outbreak, this formulation provides a practical basis for rapid assessment of whether isolation-based containment is biologically plausible, independent of whether case detection is symptom-driven or test-driven. ^52^

A central implication of the transmission waiting time framework is that intrinsic controllability cannot be inferred from *R*_0_ alone. Applying the framework across diverse directly transmitted pathogens reveals that pathogens with similar *R*_0_ may nonetheless differ markedly in containment feasibility because of differences in transmission timing. Pathogens characterized by long generation intervals, such as Ebola and smallpox, admit relatively wide temporal windows for intervention, consistent with historical containment successes. In contrast, pathogens such as influenza can be intrinsically difficult to contain even at modest *R*_0_, because rapid inter-generational turnover compresses the intervention window to only a few days. ^3,4,32,33,53^ The evolutionary trajectory of SARS-CoV-2 further illustrates how these timing constraints can tighten over short timescales: successive variants combined increasing transmissibility with progressively shorter transmission waiting times, pushing the critical isolation coverage toward unity and eroding the feasibility of isolation-based containment in most settings.^34–39^

The framework also provides a parsimonious operational interpretation of complex intervention systems. We show that heterogeneous components of test–trace–isolate (TTI) strategies can be summarized by two quantities: isolation coverage *P*_iso_ and the isolation-delay distribution *δ*_iso_(*τ* ). ^20–22^ Testing frequency, diagnostic sensitivity, and turnaround time primarily shift *δ*_iso_(*τ* ) earlier, whereas tracing efficiency and adherence primarily increase *P*_iso_. Our simulation analysis illustrates how incremental improvements along these axes move realistic TTI programs through a low-dimensional delay–coverage landscape toward the containment boundary, clarifying why isolated improvements are often insufficient: for fast-transmitting pathogens, containment typically requires coordinated gains that simultaneously compress delays and increase coverage. This representation also accommodates additional interventions—such as masking, ventilation, distancing, or temporary closures—through their effects on contact rates and interaction structure, which effectively modify the realized infectiousness profile and shift the containment frontier.^54^

We further show that transmission heterogeneity modifies controllability primarily through temporal redistribution of infectiousness, rather than through variation in individual reproduction numbers alone. Under uniform isolation, heterogeneity in individual transmissibility that is independent of infectiousness timing integrates out of the population-average *R*_*e*_ and therefore does not alter the containment condition relative to the homogeneous benchmark. By contrast, heterogeneity in infectiousness timing can either tighten or relax isolation-speed requirements depending on its coupling to transmissibility.^42^ When transmissibility is weakly coupled to mean generation interval, greater dispersion increases the contribution of early transmission from short-duration infections, tightening speed requirements. When transmissibility increases with duration, these effects are attenuated—and can even reverse—because transmission is concentrated among longer-lasting infections that are more interceptable by interventions. Taken together, these results suggest that homogeneous predictions provide a robust benchmark and, under empirically plausible positive correlations between infectiousness timing and overall shedding intensity, often a conservative bound on the speed required for containment.

Several limitations highlight priorities for future work. First, our analytical results rely on simplified mixing assumptions and uniform intervention effects, whereas real outbreaks occurs on structured contact networks with setting-specific risks and correlated behaviors that can shape both transmission timing and intervention performance. ^12,51,52,54^ Extending the framework to incorporate contact structure and heterogeneity in intervention uptake—while retaining a tractable mapping to delay and coverage—would improve realism and interpretability. Second, we treat *R*_0_ and *g*(*τ* ) as fixed pathogen characteristics, although both vary across contexts due to behavior, immunity, and non-pharmaceutical interventions. Time-varying natural history and immune landscapes may therefore shift *w*(*τ* ) and the controllability over the course of an epidemic. Third, we do not explicitly model feedbacks between control and pathogen evolution; early emergence can involve rapid adaptation that alters transmissibility, immune escape, or within-host kinetics, thereby changing the relevant transmission timescale.^9^ Integrating evolutionary dynamics with operational constraints remains an important direction for prospective risk assessment. Finally, our intervention model is restricted to single-step contact tracing. Iterative tracing, particularly when accelerated by digital technologies and paired with rapid molecular diagnostics, warrants further exploration as a means to substantially strengthen outbreak control.

In conclusion, the transmission waiting time provides a symptom-independent timescale for outbreak controllability in the modern era of molecular diagnostics. By anchoring containment to the earliest transmission event, the framework yields interpretable benchmarks for the minimum speed and reach of isolation required for control, enables principled comparisons of intrinsic controllability across pathogens, and maps complex intervention systems onto a delay–coverage landscape that directly reflects operational levers. These features make the framework well suited to guide early-stage decision-making, prioritize investments in testing and tracing performance, and clarify when isolation-based containment is feasible versus when mitigation becomes the only realistic option.

## Methods

### 1 Analytical expression for the transmission waiting time under homogeneous mixing

To quantify the timing of the earliest secondary transmission event generated by an infected individual, we adopt a survival-analysis framework and define the transmission waiting time as the interval between the time of primary infection and the occurrence of the first onward transmission event.^55^ Under homogeneous mixing, individual infectiousness over infection age *τ* is fully characterized by the basic reproduction number *R*_0_ and the generation-interval distribution *g*(*τ* ).

We assume secondary transmission events arise from a non-homogeneous Poisson process with hazard

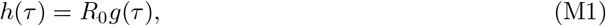

where *g*(*τ* ) is a probability density function satisfying

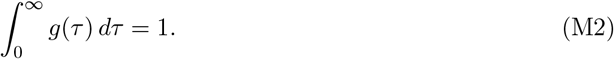

Let 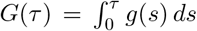 denote the corresponding cumulative distribution function. The survival function—the probability that no secondary transmission has occurred by time *τ* —is

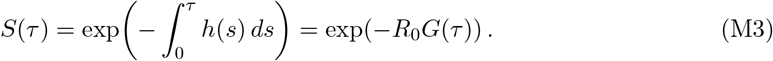

Therefore, the (defective) cumulative distribution function for the time of the first secondary transmission is

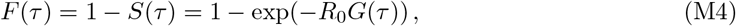

and the corresponding (defective) density is

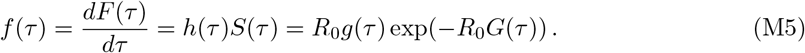

Since *S*(∞) = exp(*−R*_0_), the probability that an infection generates at least one secondary transmission is

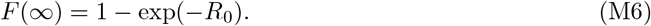

Conditioned on at least one secondary transmission occurring, the normalized transmission waiting-time density, denoted *w*(*τ* ), is

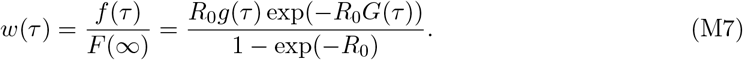

Together, *R*_0_ controls the overall intensity of transmission while *g*(*τ* ) governs the temporal profile of infectiousness; both jointly determine the distribution of the earliest transmission event.

### 2 How pathogen characteristics shape waiting time: stochastic ordering analysis

This section shows how *w*(*τ* ) shifts when (i) transmissibility increases (larger *R*_0_) or (ii) infectiousness occurs earlier (a stochastically smaller generation interval).

The conditional CDF of the transmission waiting time is obtained by integrating (M7):

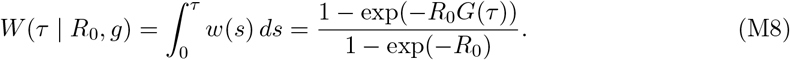

#### 2.1 Dependence on the reproduction number *R*_0_

Fix *g* (hence *G*(*τ* )). For any fixed *τ* with *G*(*τ* ) *∈* (0, 1), define

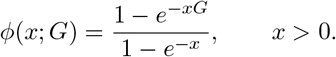

Then *W* (*τ* | *R*_0_, *g*) = *ϕ*(*R*_0_; *G*(*τ* )). One can verify that *ϕ*(*x*; *G*) is strictly increasing in *x* for each fixed *G ∈* (0, 1) (e.g., by differentiating and showing *ϕ*^*′*^(*x*; *G*) *>* 0). Therefore, if 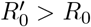, then for all *τ*,

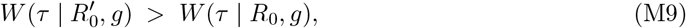

implying first-order stochastic dominance: the waiting time under 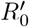 is stochastically smaller (earlier) than under *R*_0_.

#### 2.2 Dependence on the generation interval distribution

Fix *R*_0_. Suppose *g* is stochastically smaller than *g*^*′*^ in the sense that

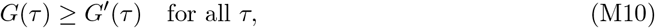

i.e., *g* concentrates more mass earlier in time. Define

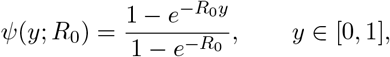

which is strictly increasing in *y*. Since *W* (*τ* | *R*_0_, *g*) = *ψ*(*G*(*τ* ); *R*_0_), condition (M10) implies

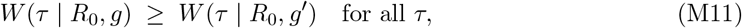

so the transmission waiting time associated with the “faster” generation interval *g* is stochastically smaller (earlier) than that associated with *g*^*′*^.

### 3 Derivation of critical intervention thresholds (homogeneous mixing case)

#### 3.1 Effective reproduction number 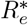 under the critical isolation timing benchmark and perfect coverage

Consider an isolation-based intervention characterized by coverage *P*_iso_ and an isolation-delay density *δ*_iso_(*τ* ) among those isolated. The probability that an infected individual remains unisolated at infection age *τ* is

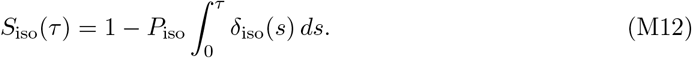

Under perfect coverage (*P*_iso_ = 1) and the critical timing benchmark 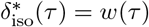, we have

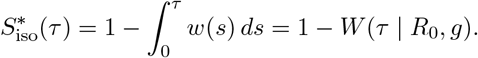

Using (M8), this becomes

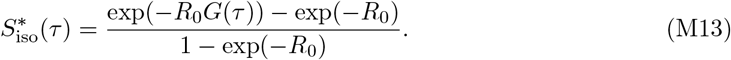

The effective reproduction number under intervention is

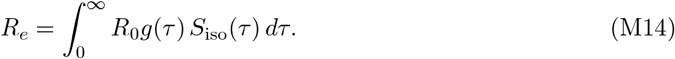

Plugging (M13) into (M14) yields

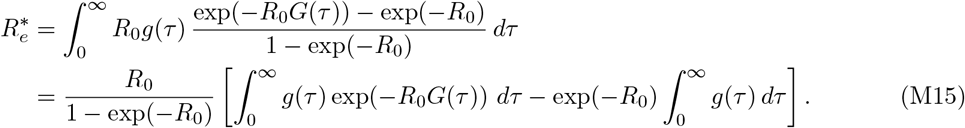

Using the substitution *u* = *G*(*τ* ) (so *du* = *g*(*τ* ) *dτ* ), the first integral becomes 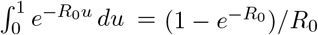, and the second integral equals 1. Therefore,

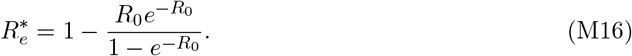

#### 3.2 Minimum required isolation coverage 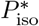 under the critical timing benchmark

Now impose the same critical timing benchmark 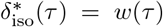 but allow imperfect coverage *P*_iso_ *<* 1. From (M12),

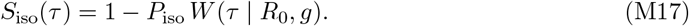

Setting the containment threshold *R*_*e*_ = 1 in (M14) gives

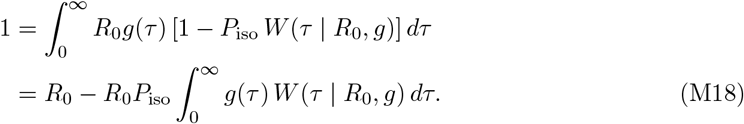

Using (M8), and substituting *u* = *G*(*τ* ),

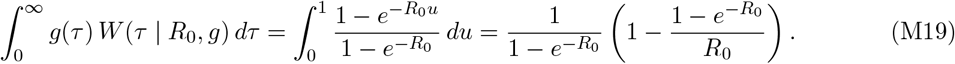

Plugging (M19) into (M18) and solving for *P*_iso_ yields

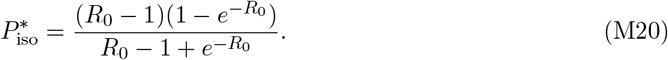

### 4 Computational framework for heterogeneity analysis

This section specifies the model and numerical procedures used to generate Figure 2. We quantify how heterogeneity in infectiousness timing reshapes controllability under a uniform isolation intervention with perfect coverage (*P*_iso_ = 100%). Notation follows the main text.

#### 4.1 Heterogeneity model for infectiousness timing and transmissibility

Each infection is characterized by an individual generation-interval distribution parameterized by its mean *µ*_*g*_. Conditional on *µ*_*g*_, we assume

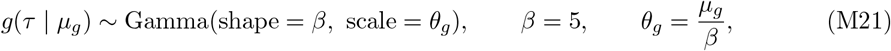

so that 𝔼 [*τ* | *µ*_*g*_] = *µ*_*g*_.

Across individuals, the mean timing parameter varies as

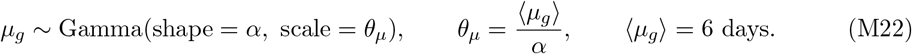

Smaller *α* corresponds to greater dispersion in infectiousness timing, while *α →* ∞ recovers the homogeneous limit *µ*_*g*_ = ⟨*µ*_*g*_⟩.

Individual transmissibility is allowed to scale with infectiousness timing via

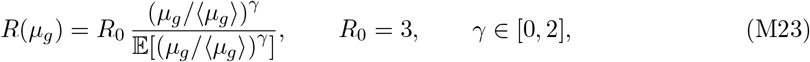

which ensures 𝔼 [*R*(*µ*_*g*_)] = *R*_0_ for all (*α, γ*).

#### 4.2 Isolation-based intervention model and homogeneous limit

We consider a uniform isolation-based intervention with perfect coverage (*P*_iso_ = 100%), parameterized by the mean isolation delay ⟨*τ*⟩_iso_. The key design feature of the intervention model is that ⟨*τ*⟩_iso_ is defined in the same waiting-time framework as pathogen transmission, ensuring a well-defined and interpretable homogeneous limit as *α →* ∞. Details of the numerical implementation is as follows:

- We first introduce an auxiliary generation-interval distribution with the same shape parameter of the individual generation interval distribution

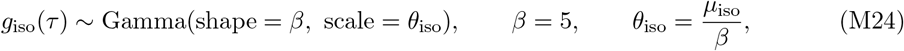

where 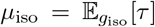 serves as a numerical control parameter that sets the characteristic timescale of isolation delay.
- We then define the isolation-delay distribution *δ*_iso_(*τ* ) as the transmission waiting time distribution induced by *g*_iso_ under a homogeneous mixing assumption with reproduction number *R*_0_ = 3:

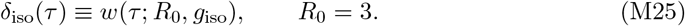

The corresponding survival function entering the transmission kernel is

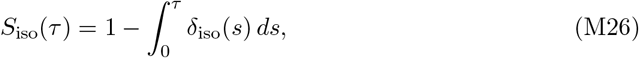

and the mean isolation delay reported in the figure is the mean of the constructed isolation delay distribution,

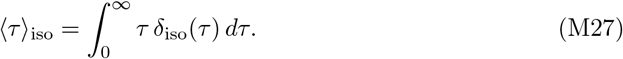

This construction ensures that the intervention is expressed in the same waiting-time language as the homogeneous mixing scenario. In particular, when infectiousness timing becomes homogeneous (*α →* ∞, *µ*_*g*_ *→* ⟨*µ*_*g*_⟩), the population transmission process is fully characterized by the homogeneous generation-interval distribution *g*(*τ* | ⟨*µ*_*g*_⟩), and the corresponding transmission waiting time reduces to *w*(*τ* ; *R*_0_, *g*(*τ* | ⟨*µ*_*g*_⟩)). Choosing *δ*_iso_ to match this distribution therefore recovers the homogeneous benchmark for controllability. Deviations in the critical isolation delay for finite *α* can thus be interpreted directly as the effect of heterogeneity relative to this homogeneous limit.

#### 4.3 Population-level 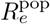, benchmark, and deviation metric

For an individual with parameters (*µ*_*g*_, *R*(*µ*_*g*_)), the expected number of secondary infections generated prior to isolation is

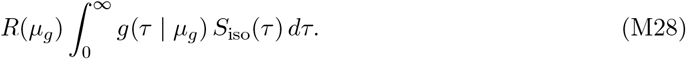

Averaging over the population distribution of *µ*_*g*_ yields

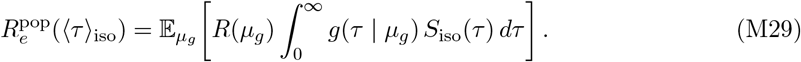

Numerically, we precompute the kernel

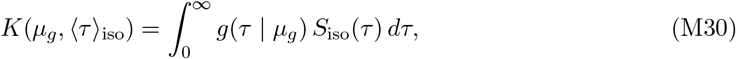

on grids *µ*_*g*_ *∈* [0.1, 100] days (1000 points) and *τ ∈* [0, 100] days with time step Δ*τ* = 0.01 days.

The dashed horizontal reference in the inset panels of Figure 2 is

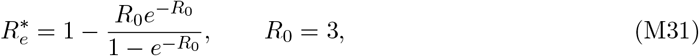

which corresponds to the homogeneous waiting-time benchmark under perfect coverage (i.e., when *δ*_iso_ matches the homogeneous transmission waiting time distribution).

For each (*α, γ*), we evaluate 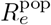 on the ⟨*τ*⟩ grid induced by the construction above and numerically identify 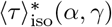 as the value satisfying

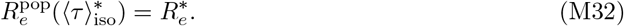

The main panel reports the deviation of this critical isolation delay relative to the homogeneous mean transmission waiting time ⟨*τ*⟩_*w*_ (computed by applying the same waiting-time transformation to the homogeneous *g*(*τ* | ⟨*µ*_*g*_⟩)):

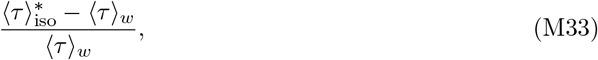

with axis tick labels displayed as percentages.

### 5 Branching process model for TTI simulations

We developed a stochastic branching-process model to evaluate the effectiveness of test–trace– isolate (TTI) strategies under realistic operational constraints. Transmission unfolds over discrete generations, with each infected individual generating a random number of secondary infections. In the absence of intervention, the basic reproduction number is fixed at *R*_0_ = 6, and generation intervals are drawn from a gamma distribution with shape parameter *k* = 5 and mean 6 days.

#### 5.1 Triggering of interventions and population-level control

The outbreak is allowed to evolve without intervention for four generations. A public-health response is then initiated at

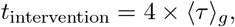

where ⟨*τ*⟩_*g*_ denotes the mean generation interval. At this time, population-level control measures— such as mobility restrictions and behavioral changes—are introduced, reducing transmissibility to a post-intervention baseline value 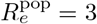 in the absence of case-based interventions (i.e., without testing, tracing, or isolation).

#### 5.2 Case-based isolation mechanisms

Following the onset of interventions, infected individuals may be isolated through three pathways:

1. **Symptom-based isolation**. Symptomatic individuals self-isolate upon symptom onset with probability 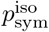. Incubation periods are drawn from a gamma distribution with shape parameter 5 and mean 4 days. If symptom onset occurs after *t*_intervention_, isolation is assumed to occur immediately at symptom onset.
2. **Routine diagnostic testing**. Individuals are screened at rate *λ*, independent of symptom status. Test sensitivity varies over the course of infection and is modeled as proportional to a proxy for infectiousness, 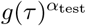, scaled by a factor *β*_test_. Following a positive test result, individuals isolate after a diagnostic turnaround delay drawn from a gamma distribution with shape parameter 5 and mean 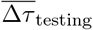 .
3. **One-step contact tracing**. Each confirmed case triggers tracing of its direct transmission offspring. The contact tracing delay is drawn from a gamma distribution with shape parameter 5 and mean 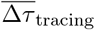 . Traced contacts isolate immediately upon notification, regardless of symptom status or test results.

#### 5.3 Computational implementation of intervention effects

To quantify intervention impact, we reconstruct the full transmission tree and remove any transmission events that occur after isolation of the infector, together with all downstream infections arising from those prevented events. The effective reproduction number under intervention, *R*_*e*_, is calculated as the mean number of secondary infections generated by individuals infected after *t*_intervention_.

From the simulated transmission trees, we extract three summary metrics: (i) the effective reproduction number *R*_*e*_ among post-intervention infections; (ii) the isolation coverage *P*_iso_, defined as the probability that an infected individual is isolated before generating any onward transmission; and (iii) the mean isolation delay ⟨*τ*⟩_iso_ among isolated individuals.

#### 5.4 Incremental intervention scenarios

We simulate six increasingly stringent TTI scenarios, each representing incremental improvements in operational performance. All scenarios assume 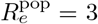, a mean generation interval of 6 days, and a mean incubation period of 4 days. Parameters are modified stepwise as follows:

1. **Scenario 1 (baseline):** 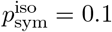, weekly testing (*λ* = 1*/*7), mean testing turnaround time of 4 days, and mean contact tracing delay of 4 days.
2. **Scenario 2:** Increase symptom-based isolation compliance to 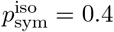.
3. **Scenario 3:** Reduce the mean contact tracing delay to 1 day.
4. **Scenario 4:** Reduce the mean testing turnaround time to 1 day.
5. **Scenario 5:** Increase test sensitivity by setting *α*_test_ = 0.5 and *β*_test_ = 0.9.
6. **Scenario 6:** Increase testing frequency to twice per week (*λ* = 2*/*7).

These scenarios illustrate the cumulative benefits of improving individual TTI components. As intervention performance improves through faster isolation, higher coverage, and more frequent and sensitive testing, the effective reproduction number declines. Under the most stringent configuration, the combined system crosses the containment threshold and achieves *R*_*e*_ *<* 1.

## Data Availability

All data produced in the present work are contained in the manuscript.

